# Clozapine-induced myocarditis and patient outcomes after drug rechallenge following myocarditis: a systematic case review

**DOI:** 10.1101/2021.09.03.21263094

**Authors:** Noah Richardson, Steven C. Greenway, Chad A. Bousman

## Abstract

Clozapine is underutilized due, in part, to concerns about rare but severe adverse drug reactions, including cardiac inflammation and injury (myocarditis). Risk factors for clozapine-induced myocarditis are limited and predictors for the successful rechallenge of clozapine after an episode of myocarditis are even more poorly understood. We conducted a systematic review, in accordance with the PRISMA recommendation, of published case reports to describe demographic and clinical characteristics of patients with clozapine-induced myocarditis and identify potential markers of clozapine rechallenge success. A total of 180 cases from 88 articles were evaluated. Male cases of clozapine-associated myocarditis were more frequently reported than female cases by a ratio of 6:1. Less than half of patients reported the presence of chest pain (35%) or flu-like symptoms (43%) but increases in troponin or C-reactive protein were present in 87% of cases. Clozapine rechallenge was carried out in 34 (2 female) cases, with successful reintroduction in 22 (2 female) cases (64.7%). Only chest pain during the initial trial was found to be significantly associated with rechallenge success (odds ratio = 6.87, 95% CI = 1.17 – 40.3). Standardized reporting of clozapine-induced myocarditis cases is needed to facilitate the identification of factors associated with successful rechallenge.

## 1. INTRODUCTION

The introduction of antipsychotic medications revolutionized the treatment of schizophrenia. However, up to 50% of patients will receive minimal or no benefit after two or more antipsychotic trials, meeting criteria for treatment-resistant schizophrenia (Howes et al., 2017; Mørup et al., 2020). For these patients, initiation of clozapine is recommended at the earliest possible opportunity (McGorry et al., 2005; NICE, 2014) and there is evidence for its superior efficacy in this difficult-to-treat population compared with other pharmacological therapies (Siskind et al., 2016). However, clozapine remains underutilized due, in part, to the risk of unpredictable and often fatal adverse drug reactions, such as myocarditis (Dvalishvili et al., 2021; Farooq et al., 2019; Singh et al., 2020).

Myocarditis involves inflammation of the heart muscle (myocardium) that typically develops within the first four weeks after commencing clozapine (Haas et al., 2007). Symptoms of myocarditis include those of heart failure and an infectious illness that when detected, will typically trigger the immediate cessation of clozapine. The incidence of clozapine-induced myocarditis is estimated to range from 3% (Ronaldson et al., 2015) to 8.5% (Reinders et al., 2004) in the context of systematic monitoring, with fatality rates ranging from 7% to 75% (Ronaldson et al., 2015). Patients with a history of clozapine-induced myocarditis have limited treatment options and, in some cases, a clozapine rechallenge is considered.

Unfortunately, our understanding of the clinical factors associated with a patient’s risk of developing or redeveloping myocarditis is currently limited and there are no protocols or guidelines for identifying patients at high risk for myocarditis before beginning clozapine treatment or for determining which patients are most suitable for clozapine rechallenge. To assist in the future development of such protocols and guidelines, we undertook a systematic review of clozapine-induced myocarditis case reports to describe the collective clinical experience with this severe adverse drug reaction and identify potential factors that may improve success of clozapine rechallenge following myocarditis.

## 2. METHODS

The systematic case review was conducted in accordance with the Preferred Reporting Items for Systematic Reviews and Meta-Analyses (PRISMA) statement (Moher et al., 2009). Case reports of clozapine-induced myocarditis were identified in PubMed, MEDLINE, and Google Scholar. All database searches were limited to peer-reviewed, published articles written in English up to January 1, 2021. The search strategy used in PubMed was: (Case reports [Publication Type]) AND ((clozapine AND myocarditis)). Similar strategies were used for other databases. Additional reports were found by manually searching the bibliographies of all identified articles. For an article to be included in the review they must have reported individual cases of patients developing myocarditis, perimyocarditis, myopericarditis, pericarditis, or suspected myocarditis while being treated with clozapine. Patients of all ages, ethnic backgrounds, and sex were included. Articles that only reported summarized data from multiple cases were excluded.

### 2.1 Data extraction

Available patient demographics, clinical information, laboratory data and details of clozapine therapy were extracted from each eligible article. Demographic information included: age, sex, and ethnicity. Ethnicity was grouped based on a standardized biogeographic grouping system (Huddart et al., 2019). Clinical factors included: smoking status (Y/N), body mass index (BMI), presence of an inflammatory illness (i.e., diseases associated with increased CRP or tropinin levels) and concomitant medication use. Medications were reviewed for known strong or moderate inhibitors of clozapine metabolism (i.e., fluvoxamine, oral contraceptives, ciprofloxacin) as well as valproic acid, given its association with increased risk for clozapine-induced myocarditis (Barnes and Paton, 2011; Ronaldson et al., 2012). Information collected regarding clozapine therapy included: starting dose (mg/day), highest dose (mg/day), duration of clozapine use (days), clozapine concentration-to-dose (C/D) ratio, and titration slope [i.e., (highest dose - lowest dose)/ (duration of titration - 1)]. Recorded signs and symptoms of clozapine-induced myocarditis included: chest pain (Y/N), flu-like symptoms (Y/N), peak temperature (degrees Celsius), peak heart rate (beats per minute, bpm), systolic blood pressure (mmHg), left ventricular ejection fraction (%), peak troponin I (ng/L), peak C-reactive protein (CRP, mg/L), peak brain natriuretic peptide (BNP, pg/mL), trough white blood cell counts (K/uL), and peak eosinophil count (K/uL). CRP and troponin levels were also examined based on monitoring thresholds proposed by Ronaldson et al. (2011). CRP levels >100 mg/L and troponin I levels greater than 2x the upper limit of normal (ULN) were noted for cases that reported these blood markers. For cases that did not report ULN values for troponin, 30 ng/L was used as the threshold. Post-myocarditis information included: cardiac biopsy performed (Y/N), infection ruled out (Y/N), and clinical outcome (fatal/nonfatal). In articles reporting rechallenge cases, additional information collected included the elapsed time from the initial clozapine trial (days) and the outcome of the rechallenge (successful/unsuccessful).

### 2.2 Statistical analysis

Extracted data were summarized using descriptive analysis techniques, including frequencies, means, standard deviations (*sd*), medians, and interquartile ranges (*IQR*). Comparison of titration slopes from eligible cases and those of recommended titration slopes in Canada (Health Canada, 2020), Australia (ACSQHC, 2012), and the United Kingdom (NHS, 2019) were performed using a one sample Student’s t-test. Demographic, clinical and laboratory factors were assessed for their association with rechallenge success using binomial logistic regression. Odds ratios and 95% confidence interval (95% CI) were calculated. An alpha threshold of 0.05 was used to determine statistical significance. If more than one factor was found to be significant, combinatorial binomial logistic regression models were tested.

## 3. RESULTS

Our search strategy resulted in a total of 104 articles and an additional 43 articles were found by handsearching bibliographies. Of these 147 articles, 32 did not report a case(s) of clozapine-induced myocarditis and another 27 articles were excluded because they only reported summarized data (i.e. individual case data not available), were not published in English, did not include myocarditis, clozapine was not used, or the full text was not available. In total, 88 articles comprising 180 cases were identified and included in our analysis (**Figure 1**). Demographic and clinical characteristics of the 180 patients are summarized in **Table 1**. Extracted data for each individual case are available in Supplementary Table S1.

**Table 1.**
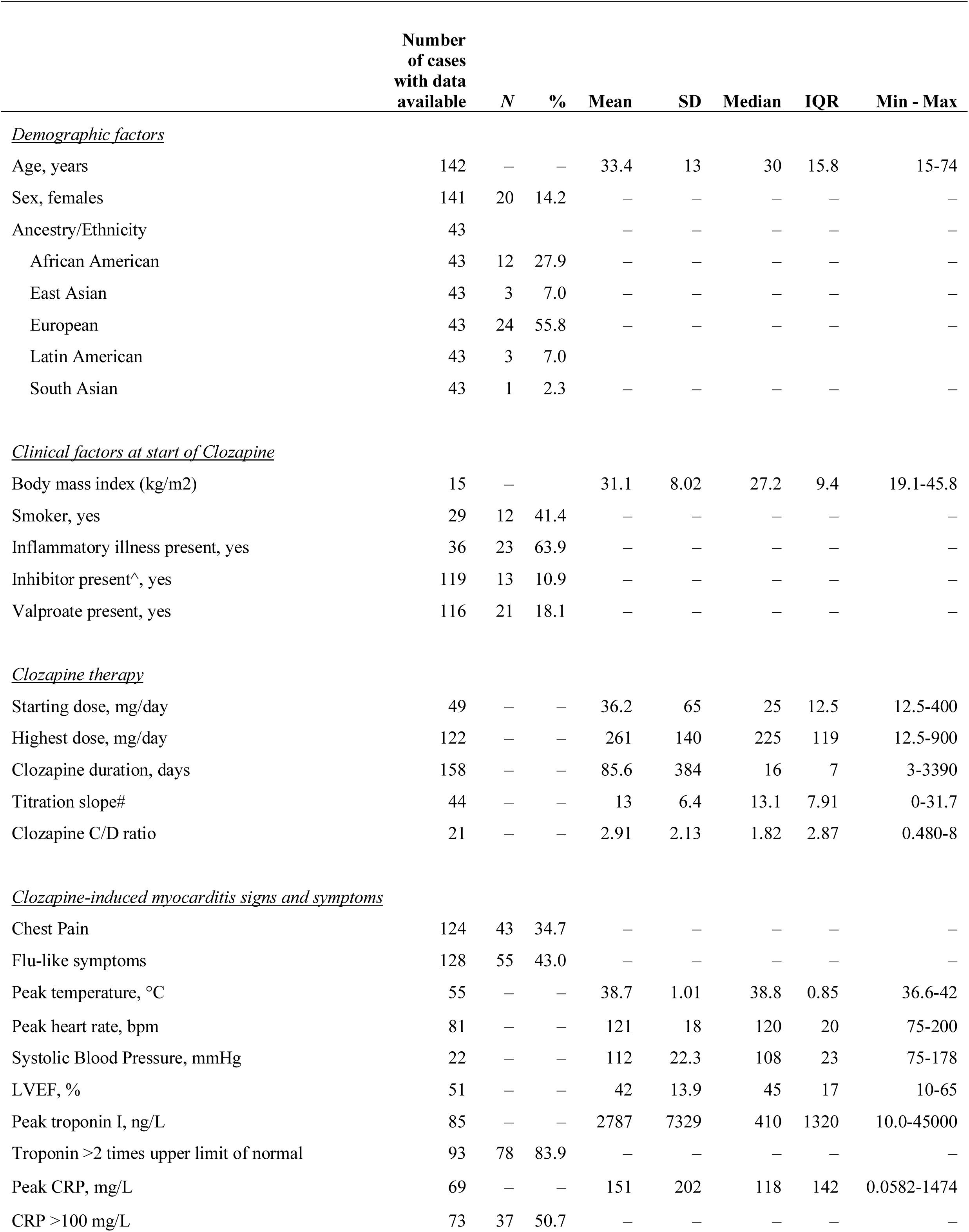
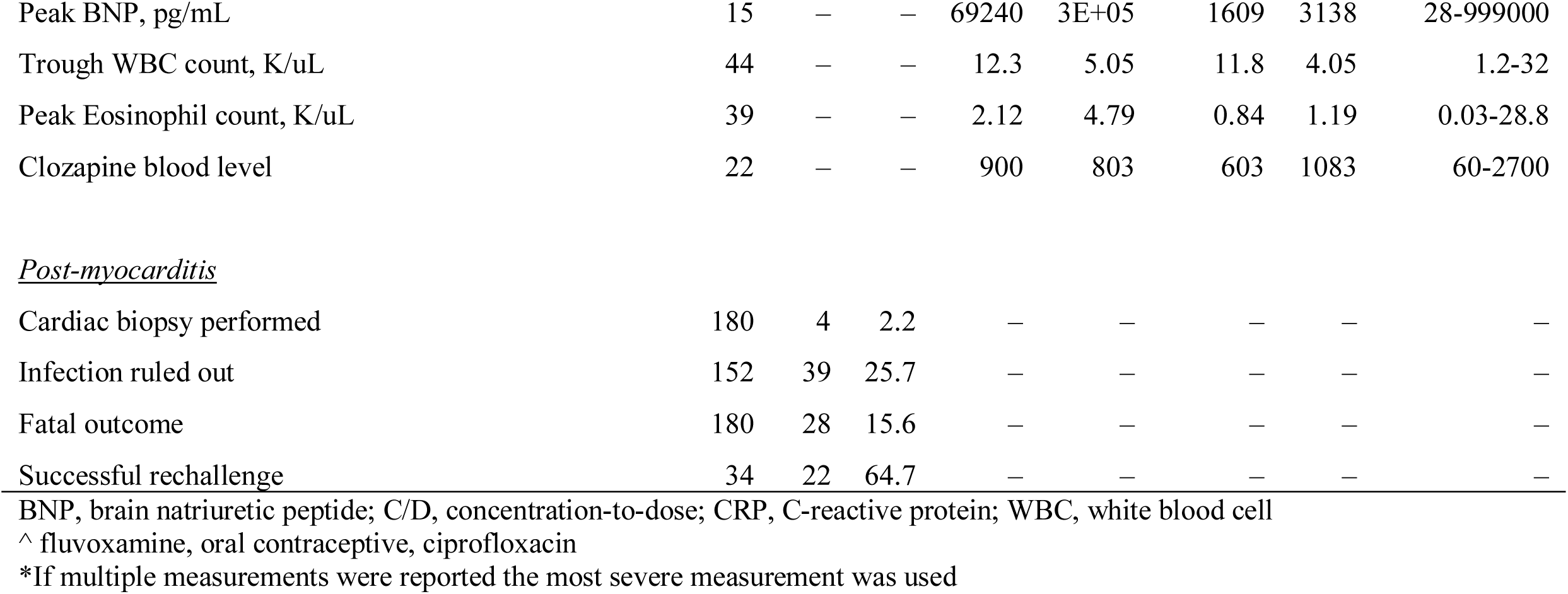
Demographic and clinical characteristics of the 180 included cases of clozapine-induced myocarditis.

|  | Number<br>of cases<br>with data<br>available | N | % | Mean | SD | Median | IQR | Min - Max |
| --- | --- | --- | --- | --- | --- | --- | --- | --- |
| *<u>Demographic factors</u>* |  |  |  |  |  |  |  |  |
| Age, years | 142 | — | — | 33.4 | 13 | 30 | 15.8 | 15-74 |
| Sex, females | 141 | 20 | 14.2 | — | — | — | — | — |
| Ancestry/Ethnicity | 43 |  |  | — | — | — | — | — |
| African American | 43 | 12 | 27.9 | — | — | — | — | — |
| East Asian | 43 | 3 | 7.0 | — | — | — | — | — |
| European | 43 | 24 | 55.8 | — | — | — | — | — |
| Latin American | 43 | 3 | 7.0 | — | — | — | — | — |
| South Asian | 43 | 1 | 2.3 | — | — | — | — | — |
| *<u>Clinical factors at start of Clozapine</u>* |  |  |  |  |  |  |  |  |
| Body mass index (kg/m2) | 15 | — | — | 31.1 | 8.02 | 27.2 | 9.4 | 19.1-45.8 |
| Smoker, yes | 29 | 12 | 41.4 | — | — | — | — | — |
| Inflammatory illness present, yes | 36 | 23 | 63.9 | — | — | — | — | — |
| Inhibitor present^, yes | 119 | 13 | 10.9 | — | — | — | — | — |
| Valproate present, yes | 116 | 21 | 18.1 | — | — | — | — | — |
| *<u>Clozapine therapy</u>* |  |  |  |  |  |  |  |  |
| Starting dose, mg/day | 49 | — | — | 36.2 | 65 | 25 | 12.5 | 12.5-400 |
| Highest dose, mg/day | 122 | — | — | 261 | 140 | 225 | 119 | 12.5-900 |
| Clozapine duration, days | 158 | — | — | 85.6 | 384 | 16 | 7 | 3-3390 |
| Titration slope# | 44 | — | — | 13 | 6.4 | 13.1 | 7.91 | 0-31.7 |
| Clozapine C/D ratio | 21 | — | — | 2.91 | 2.13 | 1.82 | 2.87 | 0.480-8 |
| *<u>Clozapine-induced myocarditis signs and symptoms</u>* |  |  |  |  |  |  |  |  |
| Chest Pain | 124 | 43 | 34.7 | — | — | — | — | — |
| Flu-like symptoms | 128 | 55 | 43.0 | — | — | — | — | — |
| Peak temperature, °C | 55 | — | — | 38.7 | 1.01 | 38.8 | 0.85 | 36.6-42 |
| Peak heart rate, bpm | 81 | — | — | 121 | 18 | 120 | 20 | 75-200 |
| Systolic Blood Pressure, mmHg | 22 | — | — | 112 | 22.3 | 108 | 23 | 75-178 |
| LVEF, % | 51 | — | — | 42 | 13.9 | 45 | 17 | 10-65 |
| Peak troponin I, ng/L | 85 | — | — | 2787 | 7329 | 410 | 1320 | 10.0-45000 |
| Troponin >2 times upper limit of normal | 93 | 78 | 83.9 | — | — | — | — | — |
| Peak CRP, mg/L | 69 | — | — | 151 | 202 | 118 | 142 | 0.0582-1474 |
| CRP >100 mg/L | 73 | 37 | 50.7 | — | — | — | — | — |
| Peak BNP, pg/mL | 15 | – | – | 69240 | 3E+05 | 1609 | 3138 | 28-999000 |
| Trough WBC count, K/uL | 44 | – | – | 12.3 | 5.05 | 11.8 | 4.05 | 1.2-32 |
| Peak Eosinophil count, K/uL | 39 | – | – | 2.12 | 4.79 | 0.84 | 1.19 | 0.03-28.8 |
| Clozapine blood level | 22 | – | – | 900 | 803 | 603 | 1083 | 60-2700 |
| Post-myocarditis |  |  |  |  |  |  |  |  |
| Cardiac biopsy performed | 180 | 4 | 2.2 | – | – | – | – | – |
| Infection ruled out | 152 | 39 | 25.7 | – | – | – | – | – |
| Fatal outcome | 180 | 28 | 15.6 | – | – | – | – | – |
| Successful rechallenge | 34 | 22 | 64.7 | – | – | – | – | – |

BNP, brain natriuretic peptide; C/D, concentration-to-dose; CRP, C-reactive protein; WBC, white blood cell
^ fluvoxamine, oral contraceptive, ciprofloxacin
\*If multiple measurements were reported the most severe measurement was used

**Figure 1.**
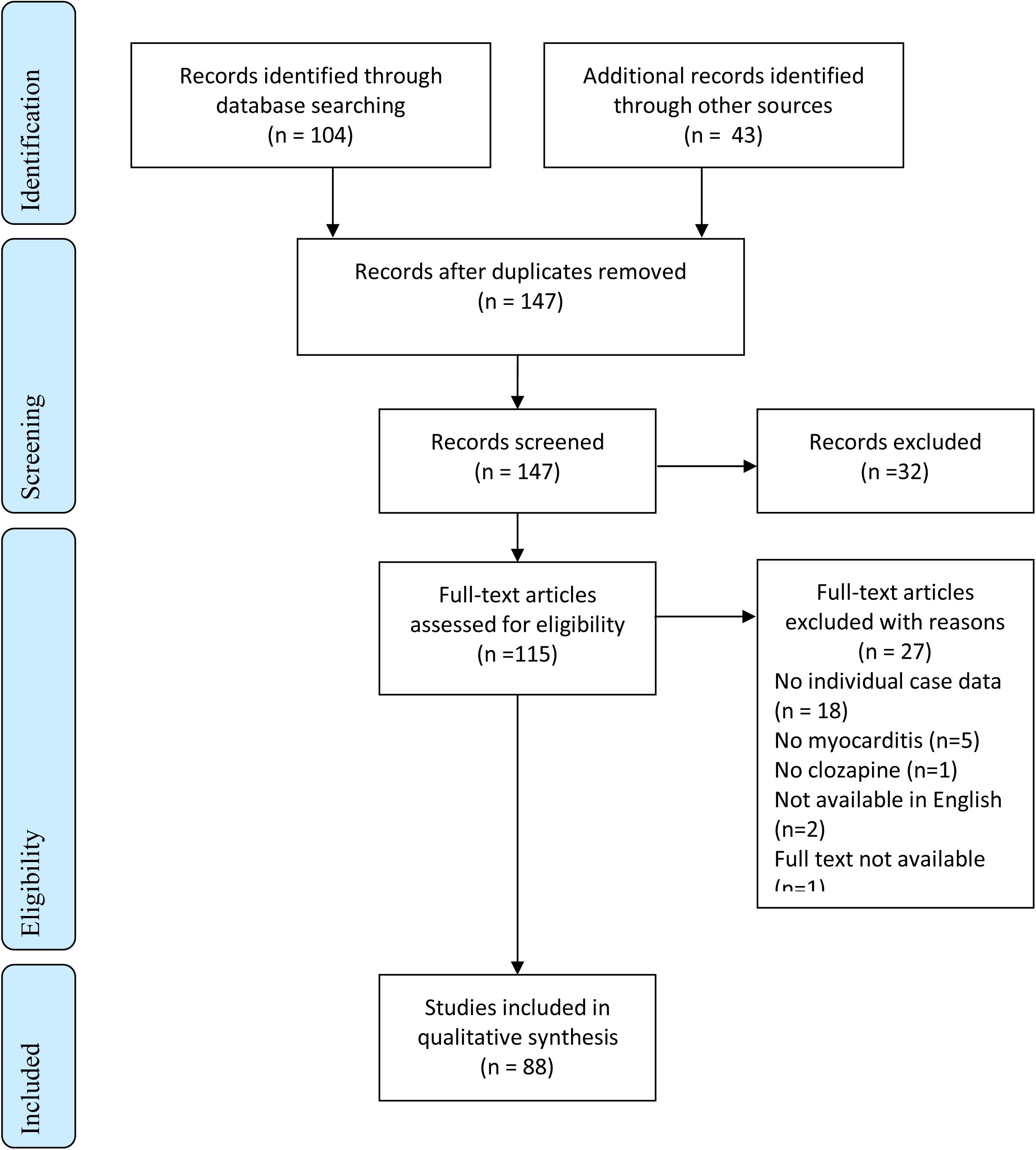
PRISMA flowchart.

### 3.1 Demographic characteristics

The mean age of cases was 33 ± 13 years old. Male cases were more frequently reported than female cases by a ratio of 6:1 and male cases were younger than female cases (males: mean age = 32 ± 12.5 years vs. females: 41.5 ± 15.3 years, *P*=0.003). Of the 43 cases with ancestry/ethnicity information available, most were of European/Caucasian background (55.8%).

### 3.2 Clinical factors at the start of clozapine

Smoking status, BMI, and the presence of an inflammatory illness were only reported in 16% (*n*=29), 8% (*n*=15), and 20% (*n*=36) of the identified cases, respectively. Among cases with this information available, 41% (12/29) were smokers and on average were obese (mean BMI = 31 ± 8 kg/m2). The presence of an inflammatory illness at the commencement of clozapine was reported in 64% (23/36) of these cases. A total of 70 different concomitant medications were reported in 118 cases. The most commonly co-prescribed medications were valproic acid (17.8%), inhibitors of clozapine metabolism (i.e., fluvoxamine, oral contraceptives, ciprofloxacin) (10.3%), haloperidol (9.3%), quetiapine (9.3%), selective serotonin reuptake inhibitors (9.3%), risperidone (9.3%), and benztropine (7.6%) (Supplementary Table S2).

### 3.3 Clozapine therapy factors

Starting daily doses were reported for only 49/180 (27%) of cases and the average starting dose was 36 ± 65 mg/day. The highest clozapine daily dose achieved was reported for the majority of identified cases (68%) and the average maximum dose was 261 ± 140) mg/day. On average, clozapine was administered for 86 ± 384 days among the 158 cases reporting this information, although the median (IQR) was only 16 (14-21) days. Clozapine C/D ratios could be calculated or were reported in a minority (12%) of cases, with a wide range (0.48 – 8.0). Likewise, titration slope could only be calculated for 45 (25%) of the identified cases, with an average slope of 12 ± 7. As shown in **Figure 2**, the median titration slopes were significantly lower than slopes recommended by protocols used in Canada (*p* < 0.001), United Kingdom (*p* < 0.001), or Australia (*p* = 0.022).

**Figure 2.**
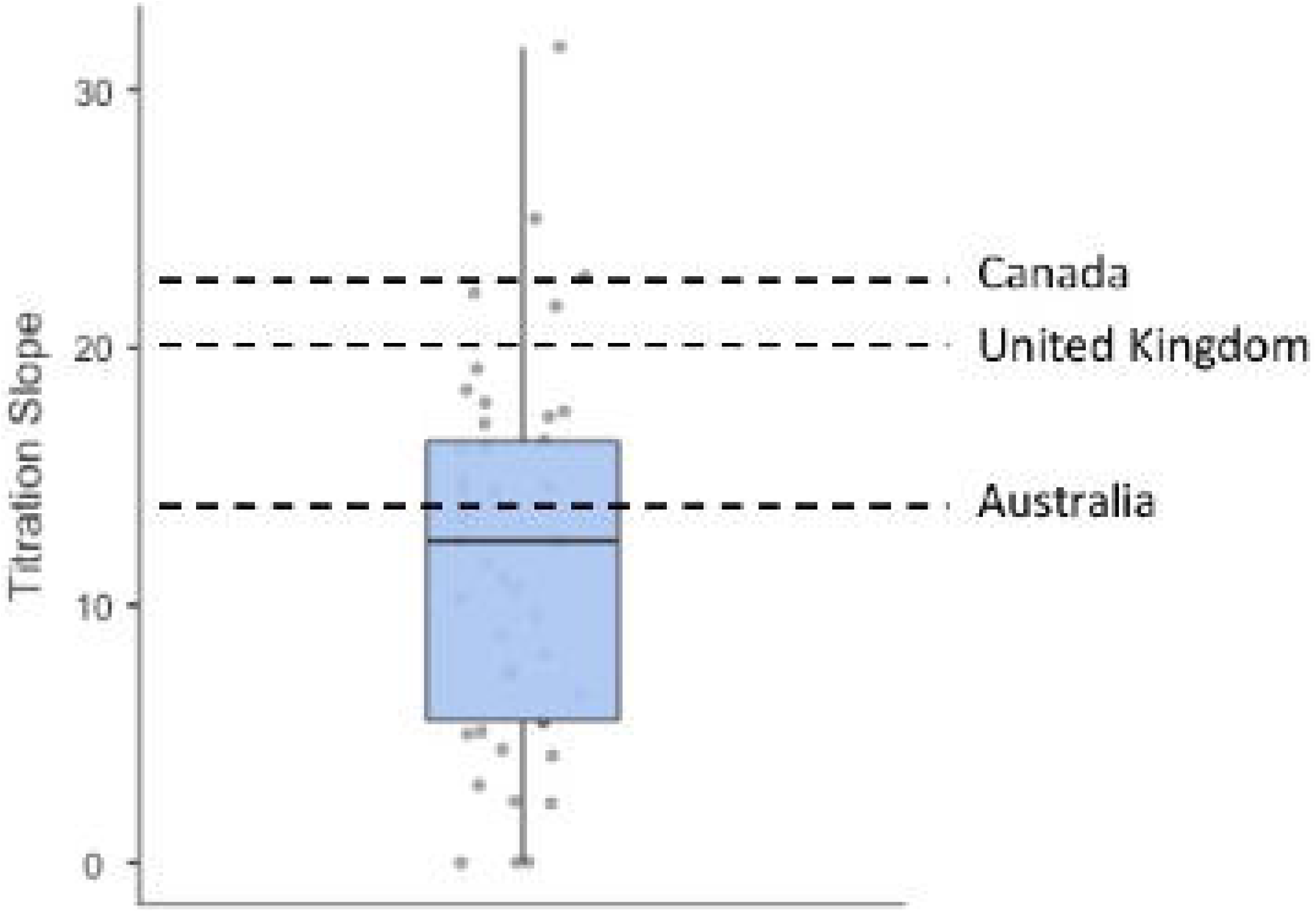
Distribution of titration slopes in reported cases of clozapine-induced myocarditis. Dashed lines represent titration slopes recommended by clozapine dosing protocols in Canada, United Kingdom, and Australia. Titration slopes were significantly lower than slopes recommended by protocols used in Canada (*p* < 0.001), United Kingdom (*p* < 0.001), or Australia (*p* = 0.022).

### 3.4 Clozapine-induced myocarditis signs and symptoms

Less than half of cases of clozapine-induced myocarditis reported the presence of chest pain (35%) or flu-like symptoms (43%). Heart rate was reported for 115 cases with 86.8% of cases having an increased heart rate (>100 bpm). As shown in Table 1, the use and reporting of laboratory tests varied considerably. The most commonly reported tests were troponin (*n*=85) and CRP (*n*=69). For troponin, 90.9% of cases had peak levels greater than the normal level (< 30 ng/L unless otherwise reported) and 83.9% had levels >2 times the upper limit of normal. For CRP, 92.2% were above the normal range (10 mg/L) and 50.7% had levels above 100 mg/L. Among the 104 cases that reported troponin or CRP levels in our review, 86.5% (*n*=90) had levels above normal. Eosinophilia (>0.5 K/uL) was present in 48.5% of the 68 cases reporting this information and among 57 cases abnormal trough white blood cell counts (5 K/uL > WBC > 10 K/uL) were reported in 52.6% of cases. Among 51 cases, the mean left ventricular ejection fraction (LVEF) was 42% (SD=14), with 36.5% having moderate or severe dysfunction (LVEF < 40%).

### 3.5 Post-myocarditis investigations and outcomes

Only four (2.2%) of the 180 cases underwent cardiac biopsy to confirm myocarditis. Among 152 cases, 25% (*n*=39) reported investigations to rule out the presence of an infection. Twenty-eight (15.6%) of all the cases we identified were fatal.

### 3.6 Clozapine rechallenge

Clozapine rechallenge was reported in 34/180 cases, among which, 22/34 cases (64.7%) were successful in reintroducing clozapine after a previous episode of myocarditis (Supplementary Table S3). The average elapsed time from clozapine discontinuation to rechallenge was 430 (*sd*=705) days. Comparisons of demographic and clinical factors between successful and unsuccessful rechallenge cases did not reveal any significant differences, with the exception of chest pain (Table 2). Cases that experienced chest pain during their initial clozapine trial had a 6.87 (95% CI = 1.171 – 40.37, *p*=0.033) greater odds of rechallenge success relative to those that did not experience chest pain during their initial clozapine trial.

**Table 2.**
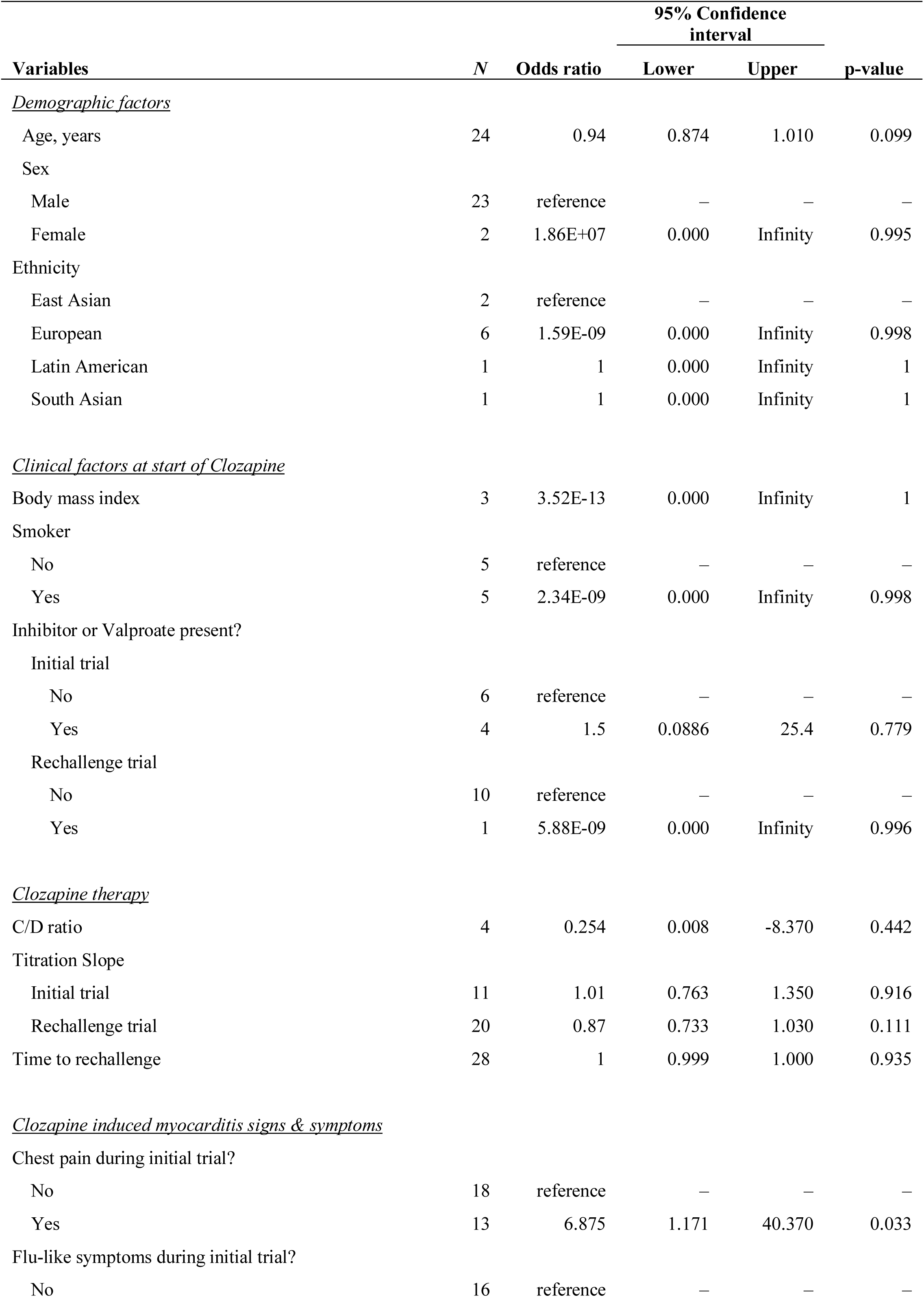
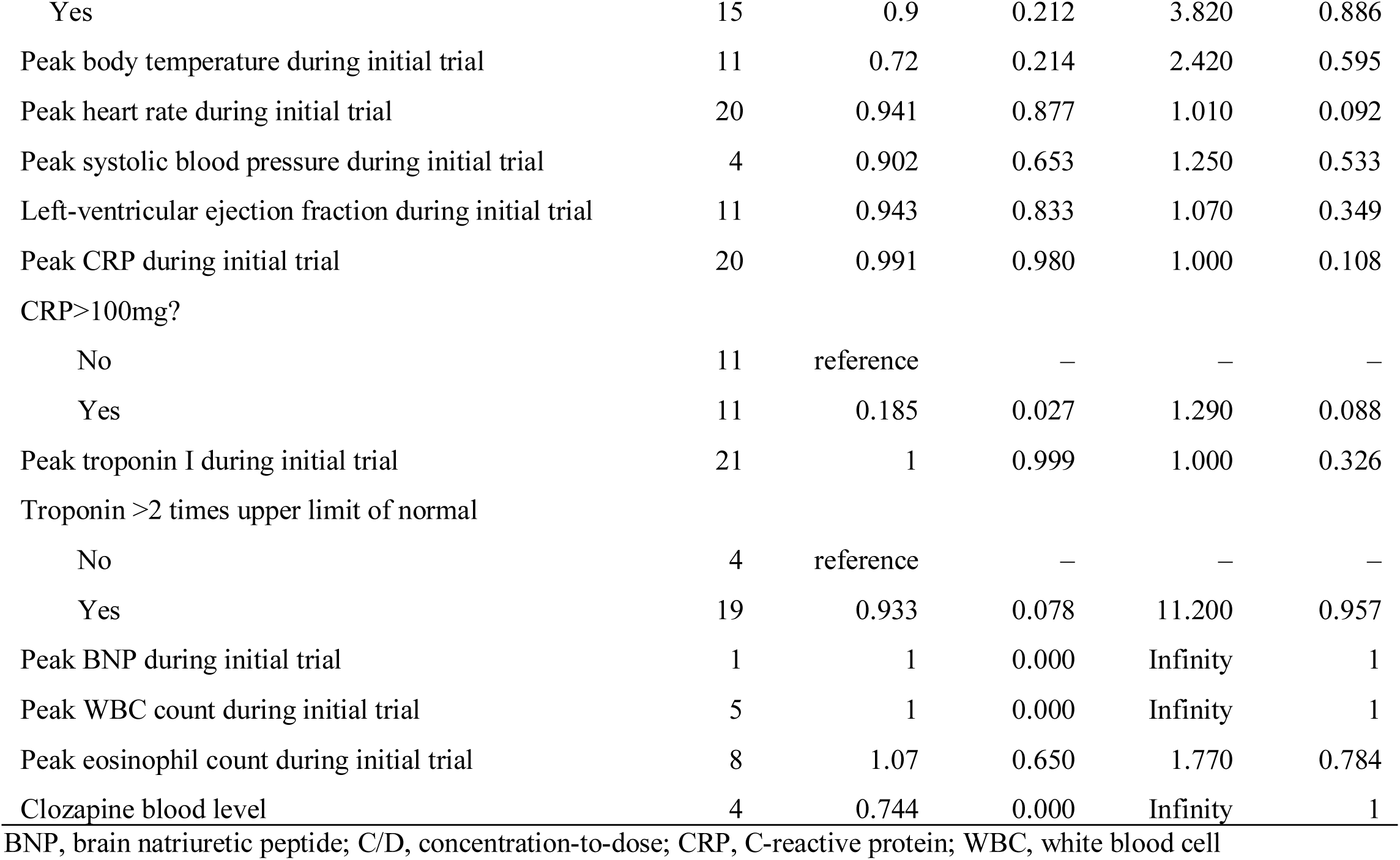
Univariate binomial logistic regression models of clozapine rechallenge success

## 4. DISCUSSION

Our systematic review of individual case reports of clozapine-induced myocarditis highlights the clinical heterogeneity of this severe adverse drug reaction and the difficulty in clinically identifying patients who are most likely to safely benefit from clozapine rechallenge. Aligned with a previous review (Bellissima et al., 2018), the clinical presentation of clozapine-induced myocarditis was extremely variable. Some of the classic signs and symptoms of myocarditis such as chest pain and flu-like symptoms were present in less than half of the cases. Other symptoms like increased heart rate were present in more cases but, unless persistent, elevated heart rate is unlikely to be a sensitive or specific enough marker to identify those at greatest risk for myocarditis but rather represents a potential signal to increase monitoring. A frequently cited and used clozapine monitoring protocol proposed by Ronaldson and colleagues (Ronaldson et al., 2011) suggests monitoring troponin I/T and CRP levels at baseline and every seven days thereafter for the first 28 days after clozapine initiation. This monitoring protocol further recommends cessation of clozapine if troponin exceeds >2 times the upper limit of normal or CRP levels are above 100 mg/L, as these thresholds were found to have 100% sensitivity for symptomatic clozapine-induced myocarditis (Ronaldson et al., 2011). Among the cases included in our review, 86.5% met one or both of these criteria, suggesting that the actual sensitivity of this monitoring protocol may be lower than originally reported or that the missed 13.5% of cases represent false-positives. The latter explanation is based on previous work that has shown that up to 65% of cases thought to be clozapine-induced myocarditis do not actually meet published criteria for the diagnosis (Winckel et al., 2015). In partial support this finding, four of the 24 potential false-positive cases were rechallenged and all but one was successful, suggesting that clozapine-treated patients who do not have either an elevated troponin or CRP may be particularly safe candidates for clozapine rechallenge.

Clozapine rechallenge following myocarditis remains controversial, is not routinely considered, and is cautioned against on product labels (HLS Therapeutics Inc., 2020). However, for some patients the clinical benefits of a rechallenge are thought to outweigh or equal the potential risk of myocarditis reoccurrence. In fact, many of the case reports included in this review described marked improvements in patient symptoms and functioning prior to the development of myocarditis that could not be sustained following clozapine discontinuation. However, guidelines for performing a clozapine rechallenge in this clinical context have not been developed, although protocols have been proposed (Shivakumar et al., 2019). Our review sought to identify potential clinical factors associated with successful clozapine rechallenge to inform future guideline development. Chest pain during the initial clozapine trial was the only identified factor for rechallenge success but the biological plausibility and clinical utility of this factor will require further investigation. We cannot rule out that this association is a false-positive given the number of tests we performed and the modest association observed, in fact this association would not survive correction for multiple testing.

It is possible that the presence of concurrent risk factors may have led to myocarditis in some of the reported cases. For example, a recently published case of a suspected clozapine-induced myocarditis reported elevated CRP prior to commencement of the initial trial of clozapine but not prior to the rechallenge (Danilewitz et al., 2021). Modest elevations in CRP (> 25 -50 mg/L) are associated with the downregulation of hepatic and extrahepatic cytochrome P450 enzymes, such as CYP1A2, resulting in increased blood concentrations of substrate drugs, including clozapine (Hefner et al., 2016). Although not definitive, waiting for CRP levels to return to normal may have contributed to subsequent successful rechallenge. As such, rechallenge might be reasonable when the initial trial of clozapine coincided with the presence of notable inflammation (CRP > 25mg/L) or use of known moderate or strong inhibitors (e.g., fluvoxamine) of clozapine metabolism.

### 4.1 Limitations

Our findings should be interpreted with several caveats. Publication reporting standards for clozapine-induced myocarditis case reports do not exist. No two case reports in our review were identical in the information they provided. Although this is undoubtedly a result of differences in standard of care across different healthcare settings, minimum standards for reporting these cases would improve the quality of future reviews on this topic. The Consensus-based Clinical Case Reporting Guidelines (Gagnier et al., 2013) should be used as the foundation for reporting clozapine-induced myocarditis cases, with particular attention given to the demographic and clinical characteristics included in Table 1. In addition, publication bias is known to occur in case reporting. Previous work has found that there is a statistically significant gender bias against female case reports accounting for about a 10% difference between male and female cases published (Allotey et al., 2017). This publication bias may have amplified the difference between male and female cases, but it does not fully explain the difference we observed. Furthermore, publication bias likely had an impact on the rechallenge analysis. There is more incentive for clinicians to publish cases of successful rather than unsuccessful rechallenges (Shivakumar et al., 2019). Therefore, our findings may overrepresent successful rechallenges. Similarly, our review was limited to case reports published in English, resulting in the exclusion of two case reports (J et al., 2015; Vesterby et al., 1980). Finally, the lack of a ‘control’ condition in case reports hindered our ability to examine and identify factors associated with the risk of developing clozapine-induced myocarditis. Future case-control studies and meta-analysis will be better suited for this objective.

### 4.2 Conclusions

Understanding the collective clinical experience with clozapine-induced myocarditis and the factors associated with successful rechallenge will improve the safety and clinical utility of clozapine. Our findings highlight the need for standardized reporting of these clinical experiences to enable the assembly of a comprehensive clinical picture of this severe adverse drug reaction and the clinical factors robustly associated with rechallenge success. Given the lack of effective therapies for treatment-resistant schizophrenia and the enormous economic and social cost of this disorder, it is imperative that information-rich case reports continue to be published alongside more rigorous case-control and cohort studies. Doing so will facilitate safer and expanded use of this highly effective medication.

## Data Availability

Data available upon request

## Acknowledgements

None

## Funding Sources

This research did not receive any specific grant from funding agencies in the public, commercial, or not-for-profit sectors.

## Conflict of interest

CAB is the founder and a shareholder of Sequence2Script Inc. The remaining authors declare that the research was conducted in the absence of any commercial or financial relationships that could be construed as a potential conflict of interest.

## References

ACSQHC, 2012. National Adult Clozapine Titration Chart User Guide.

Allotey, P., Allotey-Reidpath, C., Reidpath, D.D., 2017. Gender bias in clinical case reports: A crosssectional study of the “big five” medical journals. PLoS One 12. https://doi.org/10.1371/journal.pone.0177386

Barnes, T.R.E., Paton, C., 2011. Antipsychotic polypharmacy in Schizophrenia: Benefits and risks. CNS Drugs 25, 383–399. https://doi.org/10.2165/11587810-000000000-00000

Bellissima, B.L., Tingle, M.D., Cicović, A., Alawami, M., Kenedi, C., 2018. A systematic review of clozapine-induced myocarditis. Int. J. Cardiol. 259, 122–129. https://doi.org/10.1016/j.ijcard.2017.12.102

Danilewitz, M., Rafizadeh, R., Bousman, C.A., 2021. Successful Clozapine Rechallenge After Suspected Clozapine-Associated Myocarditis: A Case Report. J. Clin. Psychopharmacol. 41, 218–220. https://doi.org/10.1097/JCP.0000000000001339

Dvalishvili, M., Miller, B.J., Surya, S., 2021. Comfort Level and Perceived Barriers to Clozapine Use: Survey of General Psychiatry Residents. Acad. Psychiatry. https://doi.org/10.1007/s40596-021-01468-1

Farooq, S., Choudry, A., Cohen, D., Naeem, F., Ayub, M., 2019. Barriers to using clozapine in treatment-resistant schizophrenia: systematic review. BJPsych Bull. 43, 8–16. https://doi.org/10.1192/bjb.2018.67

Gagnier, J.J., Kienle, G., Altman, D.G., Moher, D., Sox, H., Riley, D., Allaire, A., Aronson, J., Carpenter, J., Gagnier, J., Hanaway, P., Hayes, C., Jones, D., Kaszkin-Bettag, M., Kidd, M., Kiene, H., Kienle, G., Kligler, B., Knutson, L., Koch, C., Milgate, K., Mittelman, M., Oltean, H., Plotnikoff, G., Rison, R.A., Sethi, A., Shamseer, L., Smith, R., Tugwell, P., 2013. The CARE guidelines: Consensus-based clinical case reporting guideline development. BMJ Case Rep. 2013. https://doi.org/10.1136/bcr-2013-201554

Haas, S.J., Hill, R., Krum, H., Liew, D., Tonkin, A., Demos, L., Stephan, K., McNeil, J., 2007. Clozapine-associated myocarditis: A review of 116 cases of suspected myocarditis associated with the use of clozapine in Australia during 1993-2003. Drug Saf. https://doi.org/10.2165/00002018-200730010-00005

Health Canada, 2020. Clozaril product monograph [WWW Document].

Hefner, G., Shams, M.E.E., Unterecker, S., Falter, T., Hiemke, C., 2016. Inflammation and psychotropic drugs: The relationship between C-reactive protein and antipsychotic drug levels. Psychopharmacology (Berl). 233, 1695–1705. https://doi.org/10.1007/s00213-015-3976-0

HLS Therapeutics Inc., 2020. Clozaril product monograph [WWW Document]. URL http://www.hlstherapeutics.com/wp-content/uploads/monograph_pdf/HLS-Clozaril-PM-E.pdf

Howes, O.D., McCutcheon, R., Agid, O., De Bartolomeis, A., Van Beveren, N.J.M., Birnbaum, M.L., Bloomfield, M.A.P., Bressan, R.A., Buchanan, R.W., Carpenter, W.T., Castle, D.J., Citrome, L., Daskalakis, Z.J., Davidson, M., Drake, R.J., Dursun, S., Ebdrup, B.H., Elkis, H., Falkai, P., Fleischacker, W.W., Gadelha, A., Gaughran, F., Glenthøj, B.Y., Graff-Guerrero, A., Hallak, J.E.C., Honer, W.G., Kennedy, J., Kinon, B.J., Lawrie, S.M., Lee, J., Leweke, F.M., MacCabe, J.H., McNabb, C.B., Meltzer, H., Möller, H.J., Nakajima, S., Pantelis, C., Marques, T.R., Remington, G., Rossell, S.L., Russell, B.R., Siu, C.O., Suzuki, T., Sommer, I.E., Taylor, D., Thomas, N., Üçok, A., Umbricht, D., Walters, J.T.R., Kane, J., Correll, C.U., 2017. Treatment-ResistantSchizophrenia: TreatmentResponse and Resistance in Psychosis (TRRIP) Working Group Consensus Guidelines on Diagnosis and Terminology. Am. J. Psychiatry 174, 216–229. https://doi.org/10.1176/appi.ajp.2016.16050503

Huddart, R., Fohner, A.E., Whirl-Carrillo, M., Wojcik, G.L., Gignoux, C.R., Popejoy, A.B., Bustamante, C.D., Altman, R.B., Klein, T.E., 2019. Standardized Biogeographic Grouping System for Annotating Populations in Pharmacogenetic Research. Clin. Pharmacol. Ther. 105, 1256–1262. https://doi.org/10.1002/cpt.1322

J, M.M. de N., JJ, P.F., E, G.M., B, D.A., 2015. Miocarditis aguda secundaria a clozapina [Clozapine-associated myocarditis]. Med. Clin. (Barc). 145, e19–e20. https://doi.org/10.1016/J.MEDCLI.2015.01.004

McGorry, P., Killackey, E., Lambert, T., Lambert, M., Jackson, H., Codyre, D., James, N., Pantelis, C., Pirkis, J., Jones, P., Durie, M.A., McGrath, J.J., McGlashan, T., Malla, A., Farhall, J., Herman, H., Hocking, B., 2005. Royal Australian and New Zealand College of Psychiatrists clinical practice guidelines for the treatment of schizophrenia and related disorders. Aust. N. Z. J. Psychiatry 39, 1–30. https://doi.org/10.1111/j.1440-1614.2005.01516.x

Moher, D., Liberati, A., Tetzlaff, J., Altman, D.G., Group, T.P., 2009. Preferred Reporting Items for Systematic Reviews and Meta-Analyses: The PRISMA Statement. PLoS Med. 6. https://doi.org/10.1371/JOURNAL.PMED.1000097

Mørup, M.F., Kymes, S.M., Åström, D.O., 2020. A modelling approach to estimate the prevalence of treatment-resistant schizophrenia in the United States. PLoS One 15. https://doi.org/10.1371/journal.pone.0234121

NHS, 2019. Clozapine treatment guidelines.

NICE, 2014. Psychosis and schizophrenia in adults: prevention and management | Guidance | NICE [WWW Document]. URL https://www.nice.org.uk/guidance/cg178 (accessed 6.8.21).

Reinders, J., Parsonage, W., Lange, D., Potter, J.M., Plever, S., 2004. Clozapine-Related Myocarditis and Cardiomyopathy in an Australian Metropolitan Psychiatric Service. Aust. New Zeal. J. Psychiatry 38, 915–922. https://doi.org/10.1080/j.1440-1614.2004.01481.x

Ronaldson, K.J., Fitzgerald, P.B., McNeil, J.J., 2015. Clozapine-induced myocarditis, a widely overlooked adverse reaction. Acta Psychiatr. Scand. 132, 231–240. https://doi.org/10.1111/acps.12416

Ronaldson, K.J., Fitzgerald, P.B., Taylor, A.J., Topliss, D.J., McNeil, J.J., 2011. A New Monitoring Protocol for Clozapine-Induced Myocarditis Based on an Analysis of 75 Cases and 94 Controls. Aust. New Zeal. J. Psychiatry 45, 458–465. https://doi.org/10.3109/00048674.2011.572852

Ronaldson, K.J., Fitzgerald, P.B., Taylor, A.J., Topliss, D.J., Wolfe, R., McNeil, J.J., 2012. Rapid clozapine dose titration and concomitant sodium valproate increase the risk of myocarditis with clozapine: A case–control study. Schizophr. Res. 141, 173–178. https://doi.org/10.1016/j.schres.2012.08.018

Shivakumar, G., Thomas, N., Sollychin, M., Takács, A., Kolamunna, S., Melgar, P., Connally, F., Neil, C., Bousman, C., Jayaram, M., Pantelis, C., 2019. Protocol for Clozapine Rechallenge in a Case of Clozapine-Induced Myocarditis. Can. J. Psychiatry 070674371989270. https://doi.org/10.1177/0706743719892709

Singh, B., Hughes, A.J., Roerig, J.L., 2020. Comfort Level and Barriers to the Appropriate Use of Clozapine: a Preliminary Survey of US Psychiatric Residents. Acad. Psychiatry 44, 53–58. https://doi.org/10.1007/s40596-019-01134-7

Siskind, D., McCartney, L., Goldschlager, R., Kisely, S., 2016. Clozapine v. first-and second-generation antipsychotics in treatment-refractory schizophrenia: systematic review and meta-analysis. Br. J. Psychiatry 209, 385–392. https://doi.org/10.1192/bjp.bp.115.177261

Vesterby, A., Pedersen, J.H., Kaempe, B., Thomsen, N.J., 1980. Pludselig død under behandling med klozapin (Leponex) [Sudden death during treatment with clozapine (Leponex)]. Ugeskr. Laeger 142, 170–1.

Winckel, K., Siskind, D., Hollingworth, S., Wheeler, A., 2015. Clozapine-induced myocarditis: Separating the wheat from the chaff. Aust. N. Z. J. Psychiatry. https://doi.org/10.1177/0004867414554269

