## Supplementary Material for "Clozapine-induced myocarditis and patient outcomes after drug rechallenge following myocarditis: a systematic case review"

**Supplementary Materials**

| **Supplementary Table 1**. Frequency of reported concomitant medications among the 180 clozapine-induced myocarditis cases | | |
| --- | --- | --- |
| **Medication** | **n** | **%** |
| Valproic Acid | 21 | 17.80 |
| Clozapine inhibitor (yes)^1^ | 13 | 11.02 |
| SSRI | 11 | 9.32 |
| Quetiapine | 11 | 9.32 |
| Haloperidol | 11 | 9.32 |
| Risperidone | 11 | 9.32 |
| Benztropine | 9 | 7.63 |
| Olanzapine | 6 | 5.08 |
| Escitalopram | 5 | 4.24 |
| Lorazepam | 5 | 4.24 |
| Diazepam | 5 | 4.24 |
| Sertraline | 4 | 3.39 |
| Clonazepam | 3 | 2.54 |
| Amlodipine | 3 | 2.54 |
| Metformin | 3 | 2.54 |
| Venlafaxine | 3 | 2.54 |
| Thyroxine | 3 | 2.54 |
| Paliperidone | 2 | 1.69 |
| Temazepam | 2 | 1.69 |
| Biperiden | 2 | 1.69 |
| Hydrochlorothiazide | 2 | 1.69 |
| Lithium | 2 | 1.69 |
| Propanolol | 2 | 1.69 |
| Atenolol | 2 | 1.69 |
| Aripiprazole | 2 | 1.69 |
| Flupentixol | 2 | 1.69 |
| Hydroxyzine | 2 | 1.69 |
| Glycopyrrolate | 2 | 1.69 |
| Docusate | 2 | 1.69 |
| Pantoprazole | 2 | 1.69 |
| Flunitrazepam | 2 | 1.69 |
| Pravastatin | 2 | 1.69 |
| Lisinopril | 1 | 0.85 |
| Metroprolol | 1 | 0.85 |
| Promazine | 1 | 0.85 |
| Aloperidol | 1 | 0.85 |
| Delorazepam | 1 | 0.85 |
| Lactulose | 1 | 0.85 |
| Glibenclamide | 1 | 0.85 |
| Atomoxetine | 1 | 0.85 |
| Salbutamol | 1 | 0.85 |
| Ziprasidone | 1 | 0.85 |
| Bupropion | 1 | 0.85 |
| Ranitidine | 1 | 0.85 |
| Fluoxetine | 1 | 0.85 |
| Pseudoephedrine | 1 | 0.85 |
| Amisulpride | 1 | 0.85 |
| Cariprazine | 1 | 0.85 |
| Duloxetine | 1 | 0.85 |
| Omeprazole | 1 | 0.85 |
| Lidocaine | 1 | 0.85 |
| Polyethylene | 1 | 0.85 |
| Finasteride | 1 | 0.85 |
| Tamsulosine | 1 | 0.85 |
| Acetylsalicylic Acid | 1 | 0.85 |
| Zopiclone | 1 | 0.85 |
| Metoprolol | 1 | 0.85 |
| Atorvastatine | 1 | 0.85 |
| Amiloride | 1 | 0.85 |
| Propriomazine | 1 | 0.85 |
| Busperone | 1 | 0.85 |
| Alimemazine | 1 | 0.85 |
| Chlorprothixene | 1 | 0.85 |
| Perphenazine | 1 | 0.85 |
| Cyproterone acetate | 1 | 0.85 |
| Chlorpromazine | 1 | 0.85 |
| Fluphenazine | 1 | 0.85 |
| Montelukast | 1 | 0.85 |
| Loratadine | 1 | 0.85 |
| Omeprazole | 1 | 0.85 |
| Iloperidone | 1 | 0.85 |
| Lamotrigine | 1 | 0.85 |

1 = inhibitors include fluvoxamine, oral contraceptive, ciprofloxacin

| **Supplementary Table 2**. Characteristics of 34 clozapine re-challenge case | | | | | | | | | | | | |
| --- | --- | --- | --- | --- | --- | --- | --- | --- | --- | --- | --- | --- |
|  |  |  |  |  |  | **Inhibitor or VPA present (Y/N)** | |  | **Titration slope** | |  |  |
| **Study** | **Case ID** | **Age** | **Sex** | **Ethnicity** | **Smoker (Y/N)** | **Initial Trial** | **Rechallenge Trial** |  | **Initial Trial** | **Rechallenge Trial** | **Time to Rechallenge** | **Successful Outcome (Y/N)** |
| Sarathy & Alexopoulus 2017^1^ | 4A | 29 | M | – | Y | – | – |  | 14.4 | 1.6 | 730 | Y |
| Ittasakul et al. 2016^2^ | 9A | 20 | M | – | – | N | N |  | – | 8.8 | 6 | Y |
| Goodison et al. 2015^3^ | 18A | 27 | M | – | – | – | – |  | – | – | – | Y |
| Smith et al. 2015^4^ | 24A | 45 | M | – | – | N | N |  | – | – | 61 | N |
| Chow et al. 2014^5^ | 26A | 15 | M | – | – | – | – |  | 14.4 | 3.1 | 106 | Y |
| Granja-Ingram et al. 2013^6^ | 28A | 22 | M | – | – | – | – |  | 12.9 | – | – | Y |
| Ronaldson et al. 2012^7^ | 32A | – | – | – | – | – | – |  | – | – | 9 | Y |
| Ronaldson et al. 2012^7^ | 32B | – | – | – | – | – | – |  | – | 12.5 | 243 | Y |
| Ronaldson et al. 2012^7^ | 32C | – | – | – | – | – | – |  | – | 5 | 304 | Y |
| Ronaldson et al. 2012^7^ | 32D | – | – | – | – | – | – |  | – | 2 | 578 | Y |
| Ronaldson et al. 2012^7^ | 32E | – | – | – | – | – | – |  | – | 400* | 274 | N |
| Ronaldson et al. 2012^7^ | 32F | – | – | – | – | – | – |  | – | – | 11 | N |
| Ronaldson et al. 2012^7^ | 32G | – | – | – | – | – | – |  | – | 22.9 | 213 | N |
| Ronaldson et al. 2012^7^ | 32H | – | – | – | – | – | – |  | – | 18.7 | 2920 | N |
| Hassan et al. 2011^8^ | 33A | 30s | M | – | – | – | – |  | – | 6.2 | 760 | Y |
| Ronaldson et al. 2011^9^ | 37I | – | – | – | – | Y | Y |  | – | – | – | N |
| Bray et al. 2011^10^ | 38A | 43 | M | – | – | – | – |  | – | – | – | Y |
| Rosenfeld et al. 2010^11^ | 41A | 35 | M | – | – | – | – |  | – | – | 14 | Y |
| Masopust et al. 2009^12^ | 45A | 43 | M | EUR | – | – | – |  | – | – | 61 | N |
| Jayathilake et al. 2009^13^ | 47A | 42 | M | – | Y | – | – |  | – | 4.1 | – | N |
| Reid et al. 2001^14^ | 67A | 23 | M | – | – | – | – |  | 8.9 | – | 730 | Y |
| Dina et al. 2018^15^ | 80A | 58 | F | – | – | – | N |  | – | – | – | Y |
| Ngyuen et al. 2017^16^ | 85A | 18 | M | – | Y | – |  |  | – | 9.8 | 152 | Y |
| Sanader et al. 2018^17^ | 94A | 73 | M | – | – | N | N |  | – | – | 122 | N |
| Crews et al. 2010^18^ | 109A | 17 | M | – | – | – | – |  | – | – | 517 | Y |
| Otsuka et al. 2019^19^ | 120A | 20 | M | EAS | N | Y | N |  | 5.833 | 2.0 | 72 | Y |
| Noel et al. 2019^20^ | 121A | 26 | M | EUR | – | – | – |  | 4.1666667 | 4.1 | 122 | N |
| Noel et al. 2019^20^ | 121B | 25 | M | EUR | Y | – | – |  | 13.392857 | 0 | 122 | N |
| Noel et al. 2019^20^ | 121C | 20 | M | EUR | – | – | – |  | 14.423077 | 12.5 | 106 | N |
| Shivakumar et al. 2019^21^ | 126A | 34 | F | EAS | N | Y | N |  | – | 1.4 | 2555 | Y |
| Hosseini et al. 2020^22^ | 128A | 22 | M | LAT | N | N | N |  | 19.1 | 12.5 | 20 | Y |
| Griffin et al. 2021^23^ | 130A | 33 | M | EUR | – | N | N |  | 7.3 | 2.8 | 304 | Y |
| Griffin et al. 2021^23^ | 130B | 30 | M | EUR | – | Y | N |  | – | 3.5 | 365 | Y |
| Danilewitz et al. 2021^24^ | 132A | 27 | M | SAS | N | N | N |  | 5 | 1.1 | 152 | Y |
| Summary Statistics |  | 31.1 (13.9) | 2 (8.0) | 6 (60.0) | 5 (50.0) | 4 (40.0) | 1 (9.1) |  | 10.9 (4.8) | 6.7 (6.2) | 415 (696) | 22 (64.7) |
|  |  | Mean (sd) | n (%) F | n (%) EUR | n (%) Smokers | n (%) Inhibitor present | |  | mean (sd) titration slope | | Mean (sd) | n (%) Success |
| VPA, valproate;  *value removed from analysis as it represents a single dose (no titration reported). | | | | | | | | | | | | |

1.	Sarathy K, Alexopoulos C. A successful re-trial after clozapine myopericarditis. *J R Coll Physicians Edinb*. Jun 2017;47(2):146-147. doi:10.4997/JRCPE.2017.209

2.	Ittasakul P, Archer A, Kezman J, Atsariyasing W, Goldman MB. Rapid Rechallenge with Clozapine Following Pronounced Myocarditis in a Treatment-Resistant Schizophrenia Patient. *Clin Schizophr Relat Psychoses*. 2016;10(2):120-2. doi:10.3371/1935-1232-10.2.120

3.	Goodison G, Siskind D, Harcourt-Rigg C, et al. Clarifying the diagnosis of myocarditis in a patient on clozapine. *Australas Psychiatry*. Jun 2015;23(3):311-3. doi:10.1177/1039856215576419

4.	Smith L. Clozapine: Myocarditis: case report. *Reactions Weekly*. 2015;(1551):84.

5.	Chow V, Feijo I, Trieu J, Starling J, Kritharides L. Successful rechallenge of clozapine therapy following previous clozapine-induced myocarditis confirmed on cardiac MRI. *J Child Adolesc Psychopharmacol*. Mar 2014;24(2):99-101. doi:10.1089/cap.2013.0098

6.	Granja-Ingram NM, James J, Clark N, Stovall J, Heckers S. Successful re-exposure to clozapine after eosinophilia and clinically suspected myocarditis. *Braz J Psychiatry*. Mar 2013;35(1):95-6. doi:10.1016/j.rbp.2012.10.005

7.	Ronaldson KJ, Fitzgerald PB, Taylor AJ, McNeil JJ. Observations from 8 cases of clozapine rechallenge after development of myocarditis. *J Clin Psychiatry*. Feb 2012;73(2):252-4. doi:10.4088/JCP.11l07467

8.	Hassan I, Brennan A, Carroll A, Dolan M. Monitoring in clozapine rechallenge after myocarditis. *Australas Psychiatry*. Aug 2011;19(4):370-1. doi:10.3109/10398562.2011.580749

9.	Ronaldson KJ, Fitzgerald PB, Taylor AJ, Topliss DJ, McNeil JJ. Clinical course and analysis of ten fatal cases of clozapine-induced myocarditis and comparison with 66 surviving cases. *Schizophr Res*. May 2011;128(1-3):161-5. doi:10.1016/j.schres.2011.01.017

10.	Bray A, Reid R. Successful clozapine rechallenge after acute myocarditis. *Aust N Z J Psychiatry*. Jan 2011;45(1):90. doi:10.3109/00048674.2010.533365

11.	Rosenfeld AJ, Gibbs T, Ivie R, Clarke L, Merrill DB. Successful clozapine retrial after suspected myocarditis. *Am J Psychiatry*. Mar 2010;167(3):350-1. doi:10.1176/appi.ajp.2009.09081118

12.	Masopust J, Urban A, Valis M, Malý R, Tůma I, Hosák L. Repeated occurrence of clozapine-induced myocarditis in a patient with schizoaffective disorder and comorbid Parkinson's disease. *Neuro Endocrinol Lett*. Mar 2009;30(1):19-21.

13.	Jayathilake I, Singh AK. Clozapine rechallenge after myocarditis. *Australas Psychiatry*. 2009;17(5):421-2. doi:10.1080/10398560902965302

14.	Reid P, McArthur M, Pridmore S. Clozapine rechallenge after myocarditis. *Aust N Z J Psychiatry*. Apr 2001;35(2):249. doi:10.1046/j.1440-1614.2001.0884a.x

15.	Dina A, Barlis P, van Gaal W. Clozapine-Induced Myocarditis or Acute Coronary Syndrome? Optical Coherence Tomography to the Rescue. *Case Rep Cardiol*. 2018;2018:5026107. doi:10.1155/2018/5026107

16.	Nguyen B, Du C, Bastiampillai T, Dhillon R, Tibrewal P. Successful clozapine re-challenge following myocarditis. *Australas Psychiatry*. Aug 2017;25(4):385-386. doi:10.1177/1039856217707394

17.	Sanader B, Grohmann R, Grötsch P, et al. Clozapine-Induced DRESS Syndrome: A Case Series From the AMSP Multicenter Drug Safety Surveillance Project. *Pharmacopsychiatry*. Mar 2019;52(3):156-159. doi:10.1055/a-0586-8983

18.	Crews MP, Dhillon GS, MacCabe JH. Clozapine rechallenge following clozapine-induced pericarditis. *J Clin Psychiatry*. Jul 2010;71(7):959-61. doi:10.4088/JCP.09l05692yel

19.	Otsuka Y, Idemoto K, Hosoda Y, Imamura Y, Aoki T. Clozapine-induced myocarditis: Follow-up for 3.5 years after successful retrial. *J Gen Fam Med*. May 2019;20(3):114-117. doi:10.1002/jgf2.239

20.	Noël MC, Powell V, Burton L, Panda R, Remington G. Clozapine-Related Myocarditis and Rechallenge: A Case Series and Clinical Review. *J Clin Psychopharmacol*. 2019 Jul/Aug 2019;39(4):380-385. doi:10.1097/JCP.0000000000001062

21.	Shivakumar G, Thomas N, Sollychin M, et al. Protocol for Clozapine Rechallenge in a Case of Clozapine-Induced Myocarditis. *Can J Psychiatry*. Dec 2019:706743719892709. doi:10.1177/0706743719892709

22.	Hosseini SA, Skrzypcak B, Yasaei R, et al. Successful Clozapine Re-Challenge After Suspected Clozapine-Induced Myocarditis. *Am J Case Rep*. Nov 02 2020;21:e926507. doi:10.12659/AJCR.926507

23.	Griffin JM, Woznica E, Gilotra NA, Nucifora FC. Clozapine-Associated Myocarditis: A Protocol for Monitoring Upon Clozapine Initiation and Recommendations for How to Conduct a Clozapine Rechallenge. *J Clin Psychopharmacol*. 2021 Mar-Apr 01 2021;41(2):180-185. doi:10.1097/JCP.0000000000001358

24.	Danilewitz M, Rafizadeh R, Bousman C. Successful clozapine rechallenge following suspected clozapine-associated myocarditis: A case report . *Journal of Clinical Psychopharmacology*. 2020;in press
